# Community-wide annual tuberculosis screening over four years: Findings from the HPTN 071 (PopART) trial

**DOI:** 10.1101/2025.07.08.25331093

**Authors:** Thomas Gachie, Ab Schaap, Blia Yang, Ephraim Sakala, Mwelwa Phiri, Kwame Shanaube, Rory Dunbar, Ranjeeta Thomas, Sarah Fidler, Peter Bock, Peter J. Dodd, Richard Hayes, Lily Telisinghe, Sian Floyd, Helen Ayles, HPTN 071 (PopART) Study Team

**Affiliations:** London School of Hygiene & Tropical Medicine (LSHTM), London, UK; Zambart, University of Zambia School of Medicine, Lusaka, Zambia; Health Systems Trust, Cape Town, South Africa; Sheffield Centre for Health and Related Research, School of Medicine and Population Health, University of Sheffield, Sheffield UK; Imperial College London, London, United Kingdom; Desmond Tutu TB Centre, Department of Paediatrics and Child Health, Stellenbosch University, Stellenbosch, South Africa; Department of Health Policy, London School of Economics and Political Science

## Abstract

**Background:** The World Health Organization suggest that systematic tuberculosis (TB) screening may be conducted in high prevalence settings (>0.5%), though supporting evidence is limited.

**Methods:** Between January-2014 to December-2017, the HPTN 071 (PopART) cluster-randomized trial implemented universal HIV and TB testing across 21 communities in Zambia and South Africa (SA), with TB prevalence of 0.5% and 1.6%, respectively. Trained community health workers visited households annually to offer HIV testing and TB symptom screening, with sputum collection from individuals who screened positive. Diagnostic testing used Xpert-MTB/RIF or smear microscopy, and linkage to treatment was facilitated.

We analysed TB screening and diagnosis data across three rounds (R1–R3) in Zambia and R3 in SA, where complete data were available. We examined factors associated with reporting TB symptoms and being diagnosed with TB.

**Results:** The yield of newly diagnosed TB (per 100,000 persons) increased across rounds in Zambia [R1 = 81, R2 = 93, R3 = 110; p-value (trend) = 0.003] and was higher in R3 SA (380). In R3, TB yield was higher in men (Zambia: 146; SA: 543) than women (Zambia: 76; SA: 257), and among newly diagnosed HIV-positive individuals (Zambia: 541; SA: 789) compared to HIV-negative individuals (Zambia: 48; SA: 170) and self-reported HIV-positive individuals on ART (Zambia: 105; SA: 192). In Zambia R3, participants screened twice before had 38% lower odds of being diagnosed with TB compared to those screened first time [Adjusted odds ratio = 0.62, 95% CI (0.43, 0.90)].

**Conclusion:** The PopART intervention identified undiagnosed TB through systematic TB symptom screening, particularly in men and newly diagnosed HIV-positive individuals and individuals who had not previously been screened. Yield was relatively low compared to estimated TB prevalence.

**Trial registration:** ClinicalTrials.gov <u>NCT01900977</u>

## Introduction

Tuberculosis (TB) is back to being the leading infectious cause of death worldwide, since 2023 [1]. The burden of TB disease is especially high among people living with HIV (PLHIV) and men. Zambia and South Africa (SA) are both included in the World Health Organization’s (WHO) list of 30 high HIV/TB burden countries. To reduce the burden of TB disease, WHO suggests systematic screening of the general population, in settings of high prevalence (i.e. 0.5% or higher) [2]. Available evidence suggests that, if delivered with sufficient coverage and intensity, systematic community-wide TB screening could reduce the prevalence of TB at community level [3,4], and may facilitate reaching the first of the End-TB Global Plan’s 90-(90)-90 targets, which is to reach at least 90% of individuals with TB and place them on appropriate treatment [5].

In settings with high prevalence of TB disease, such as Zambia and SA, HIV/TB co-infection is common [6], and antiretroviral therapy (ART) is known to substantially reduce the risk of developing TB disease among HIV-positive individuals, improving their chances of survival [7]. Therefore, collaborative HIV/TB strategies, aimed at HIV testing and treatment, and systematic screening for TB, could lead to better TB outcomes [8]. Not much is known on the benefits of a population-wide combined TB and HIV screening strategy for TB identification and outcomes. The HPTN 071 (PopART) trial of a Universal Testing and Treatment (UTT) intervention for HIV, provided a unique opportunity to test how feasible a combined universal TB and HIV screening and treatment strategy would be in settings such as Zambia and SA [9]. The PopART intervention focused primarily on HIV, but symptom-based TB screening was embedded in the intervention package, delivered community-wide and door-to-door, and repeated approximately annually for a period of four years [9,10].

We investigate the impact of repeated community-wide TB symptom screening on the yield of newly diagnosed TB disease and present our findings from the eight Zambian and six SA intervention communities of the HPTN 071 trial. We hypothesized that, in these settings, a combined intervention including UTT for HIV and systematic TB screening would identify a considerable amount of undiagnosed TB, especially among PLHIV and among men, and that the yield of newly diagnosed TB may decrease after several years of such a strategy.

## Methods

### Intervention

Full details of the HPTN 071 (PopART) community randomized trial (CRT) and intervention have been described elsewhere.[10] Briefly, approximately one million people from 21 densely populated urban and peri-urban communities in Zambia and Western Cape province, SA, with a high burden of HIV/TB, were included [11]. A community was defined as the catchment area of a government health facility with existing HIV care, ART provision and TB treatment services. The 21 communities (12 communities in Zambia and 9 in SA) were formed into seven matched triplets (four in Zambia and three in SA) and randomly allocated to three study arms (Arms A, B and C). Arm A received the full PopART intervention including immediate ART initiation irrespective of CD4 count. Arm B received the full PopART intervention with ART initiation according to the then current national treatment guidelines, while Arm C, the control arm, continued to receive the existing standard of care including ART initiation according to the then current national treatment guidelines.

The PopART intervention, delivered to approximately 600,000 people in Arms A and B, was a combined prevention package with several components delivered door-to-door by specially trained community health workers referred to as community HIV-care providers (CHiPs) who worked in pairs, with each pair serving a “zone” consisting of approximately 500 households. This package included annual universal home-based HIV testing and counselling; referral of HIV-positive individuals to care at the local government health facility in their communities; follow-up of HIV-positive individuals to support linkage to HIV care and retention on ART as well as various other services described elsewhere [11,12]. The PopART intervention also included universal TB symptom screening and sputum collection by CHiPs followed by referral of those diagnosed with TB to the local government health facility for treatment and this forms the basis of this manuscript.

Before the start of the trial, research assistants (RAs) conducted a census across the 14 communities and listed 96,847 and 55,853 households in Zambia and SA respectively. In three intervention rounds (R1-3), each lasting an average of 15 months, the CHiPs delivered the PopART intervention by visiting all households listed by the RAs in their assigned zone, explaining the intervention and asked permission to enumerate all household members.

### CHiPs procedures

In R1 (January-2014 to June-2015) the CHiPs aimed to contact all individuals aged 18 years and older at least once and offer them the intervention. In R2 (June 2015 to October 2016) and R3 (October 2016 to December 2017) the CHiPs expanded their target group to include individuals aged 15 years and older. CHiPs made appointments and repeated attempts to contact those absent. During R2 and R3, new strategies were implemented to increase the participation in the intervention [13].

All participants were offered the HIV prevention package and screened for TB symptoms, (i.e. cough for ≥2 weeks, drenching night sweats or unintentional loss of weight of ≥1.5 kilograms in the last month), excluding those who self-reported being on TB treatment at the time of the household visit. Two sputum samples were collected one hour apart from those who screened positive for symptoms (i.e. reported having any one of the TB symptoms) and were transported to the local government health facility for diagnosis. In Zambia, smear microscopy for HIV negative participants (at health centres) and GeneXpert for HIV positive participants or those with unknown HIV status (at district laboratories) were done, following national guidelines. In SA, all participants were tested using GeneXpert by the National Health Laboratory Service.

Results were recorded in the TB laboratory registers and presumptive TB registers, and the CHiPs delivered the test results to participants. For those newly diagnosed with TB disease, the CHiPs made efforts to try and link them to care and followed up with the clinics and participants to ensure that they had started TB treatment. Further details of CHiPs procedures are presented elsewhere [12]. All data were captured electronically on an electronic data capture device which then populated a CHiPs intervention database.

### Outcomes and explanatory variables

The two main TB outcomes were; prevalence of TB symptoms and yield of newly diagnosed TB disease, defined as the number newly diagnosed with TB per 100,000 persons screened.

In addition to the two main outcomes, we also investigated three additional outcomes representing key steps in the TB testing and treatment cascade. One, number and proportion of participants who participated among those enumerated and eligible to participate (≥18 years old for R1 and ≥15 years old for R2 and R3); Two, number and proportion of participants who self-reported being currently on TB treatment; Three, number and proportion of newly diagnosed TB disease started TB treatment.

The explanatory variables included: country (Zambia; SA), community/triplet, age-group (15-17; 18-19; 20-24; 25-29; 30-39; 40-49; ≥50 years), sex (male; female), HIV status (tested HIV negative by CHiP; self-reported HIV positive & not on ART; self-reported HIV positive & on ART; tested HIV positive by CHiP; not-tested/ indeterminate), household TB contact (yes; no), mobility in R3 only (previously resident; moved from within community; moved from outside community), and the number of times previously screened as part of the PopART intervention in R2 and R3 Zambia only (first time screening; screened once before; screened twice before).

Age group categories were informed by prior studies conducted in these settings and the epidemiology of TB and HIV [14]. However, due to data sparsity, broader age categories (15–29, 30– 39, 40–49, ≥50 years) were used for the yield of newly diagnosed TB disease outcome.

### Statistical analysis

Analyses were done separately for each country and round of intervention, with SA analysis focusing on R3 only due to previously reported data collection challenges in R1 and R2 [11]. Comparisons between rounds used Zambia data, and comparisons between countries used R3 data only.

Frequencies and percentages of the outcomes representing key steps in the TB testing-and-treatment cascade, stratified by sex, age-group and HIV status, were tabulated for each round, arm and community. For every round, we calculated the prevalence of TB symptoms and yield of newly diagnosed TB disease, with 95% confidence intervals (CIs) based on the binomial distribution. To elucidate differences in prevalence of TB symptoms and yield of TB disease by sex, age-group and HIV status, we generated proportions and binomial CIs for each subgroup, and presented these graphically by round.

A logistic regression model was fitted to identify individual and household characteristics associated with the prevalence of self-reported TB symptoms and the yield of newly diagnosed TB disease. To adjust for geographical variation across the 14 intervention communities, community fixed effects were included in all models. Since the dataset encompassed essentially the entire population of these communities rather than a random sample, multilevel (hierarchical) modelling was not necessary. Binomial variation at the individual level was the only source of random variation considered in the analysis, and standard methods for proportions and logistic regression were therefore appropriate. A forward stepwise model-building strategy (p ≤ 0.1) was applied to identify risk factors for the self-reported TB symptoms outcome. Variables identified in this model were then included as covariates in the analysis of the yield of newly diagnosed TB disease.

Analysis was done using Stata (Version 18.0) and graphs were generated in R (Version 4.2.2).

### Ethical Considerations

The study was approved by the ethics committees of the London School of Hygiene & Tropical Medicine, the University of Zambia, and Stellenbosch University. Additional Institutional Review Board approvals were given for including those aged 15 years and older in later rounds. Individuals gave informed verbal consent to participate in the intervention and informed written or witnessed consent for HIV testing.

## Results

### Enumeration and participation

In Zambia, 444,462, 409,618, 423,387 individuals were listed across R1-3 and 193,579 were listed in R3 SA. In Zambia, 56.8% (252,329), 61.8% (253,168) and 62.2% (263,382) across R1-3 and 69.4% (134,367) in R3 SA were eligible to participate in the overall CHiP intervention. Proportion eligible to participate was slightly higher among females compared to males in R1-3 Zambia and in R3 SA (Fig 1, S1 Table).

**Fig 1:**
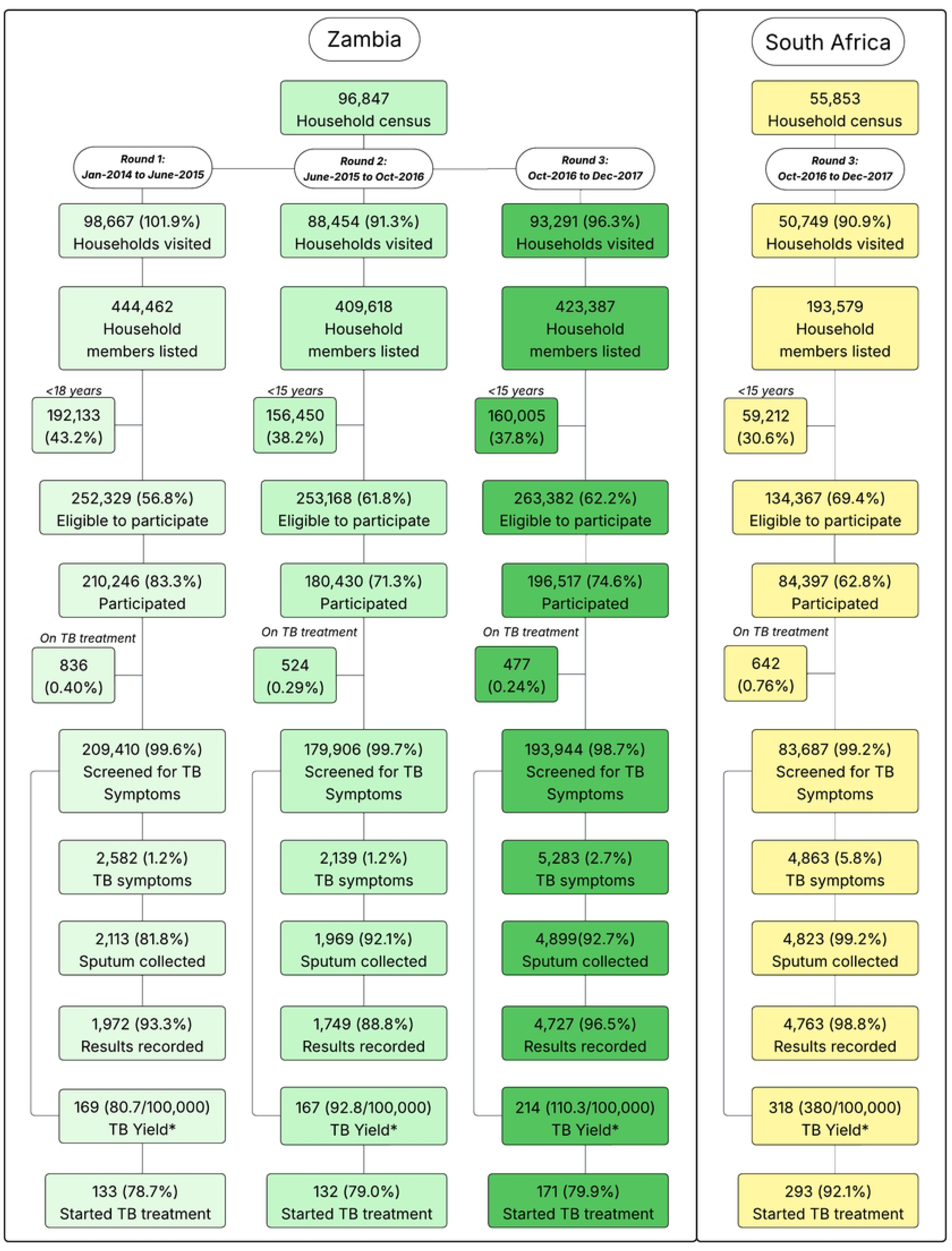
Enumeration, participation and Tuberculosis outcomes for R1-3 in Zambia and R3 in South Africa. *TB yield (per 100,000 people screened) denominator is the number screened for TB symptoms; R1-3 = Round 1-3 (January 2014 – December 2017); R3 = Round 3 (October 2016 – December 2017).

Overall, participation was relatively higher in R1 (83.3%, n = 210,246) compared to R2 (71.3%, n = 180,430) and R3 (74.6%, n = 196,517) in Zambia, and higher in Zambia R3 compared to SA R3 (62.8%, n = 84,397). Participation was higher among females compared to males and relatively similar across age-groups in R1-3 Zambia and in R3 SA (Fig 1, S1 Fig, S2 Table).

### Prevalence of self-reported TB symptoms and factors associated with having TB symptoms

Approximately 99% of participants across R1-3 Zambia (i.e. R1 = (99.6%, n = 209,410), R2 = (99.7%, n= 179,906) and R3 = (98.7%, n =193,944)) and R3 SA (99.2%, n = 83,687) were screened for TB symptoms. At the time of the CHiP’s visit 836 (0.40%), 524 (0.29%) and 477 (0.24%) participants across R1-3 in Zambia self-reported being on TB treatment and so were excluded from screening. In R3 in SA sites 642 (0.8%) self-reported being on TB treatment (Fig 1).

The prevalence of TB symptoms in Zambia was similar across R1 (1.2%, n = 2,582) and R2 (1.2%, n = 2,139) and increased in R3 (2.7%, n = 5,283), but was higher in R3 SA (5.8%, n=4863) compared to R3 Zambia (Fig 2a). This pattern was consistent across sex and age-groups, and across communities (S2-S4 Figs).

**Fig 2:**
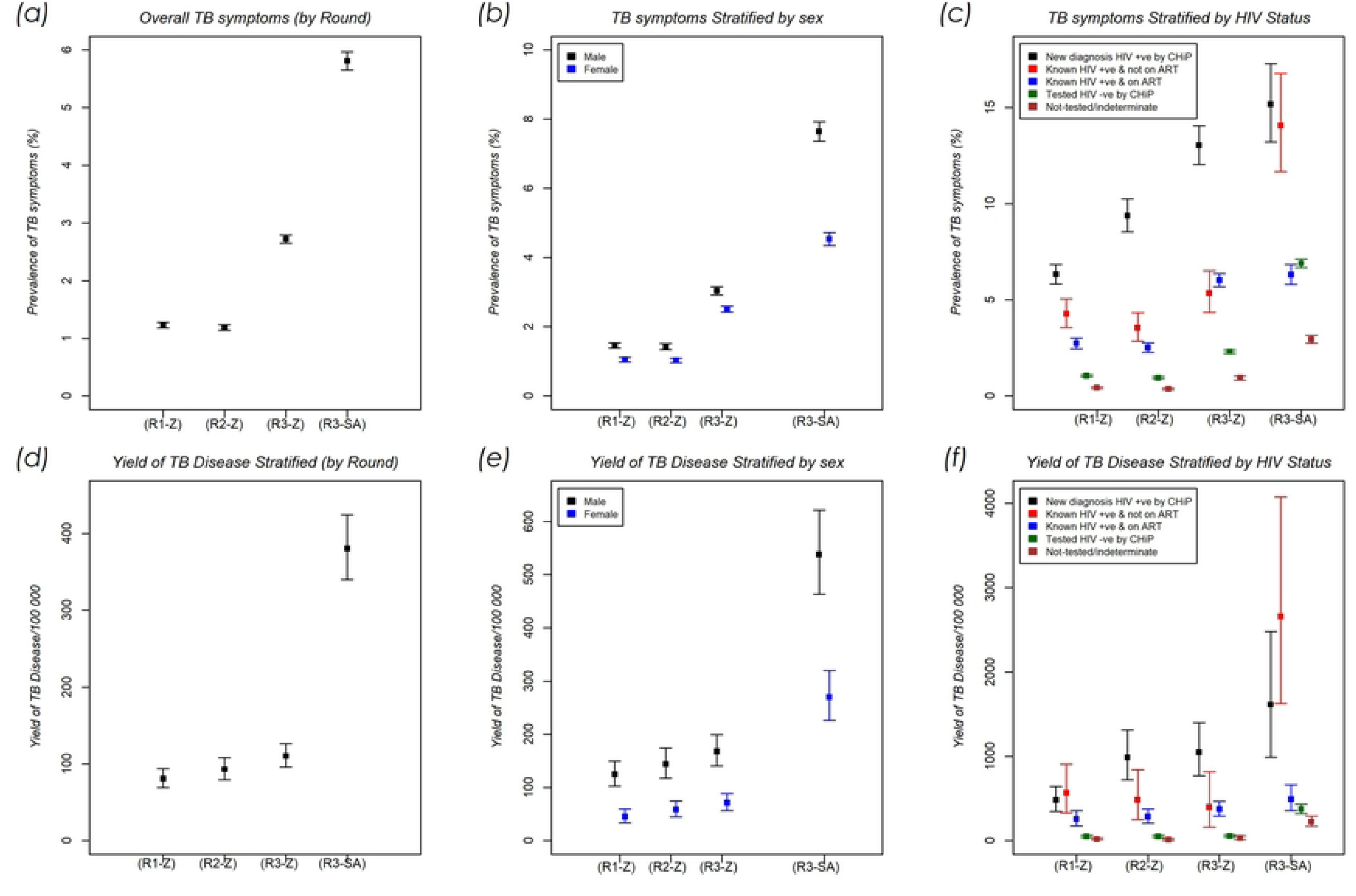
Patterns of the prevalence of TB symptoms and the yield of newly diagnosed TB Disease. Overall and stratified by sex and HIV status across R1-3 in Zambia and R3 South Africa. Note: R = Round (R1 = January 2014 to June 2015, R2 = June 2015 to October 2016, R3 = October 2016 to December 2017); SR = Self-reported; +ve = positive; −ve = negative

Across both countries, community of residence, age, sex, HIV status, household TB contact and mobility were associated with having TB symptoms (Table 1).

**Table 1:** Factors associated with having TB symptoms in Round 3 (October 2016 to December 2017) of the intervention in Zambia and South Africa.

|  | Zambia |  |  |  |  |  | South Africa |  |  |  |  |
| --- | --- | --- | --- | --- | --- | --- | --- | --- | --- | --- | --- |
|  | Descriptive analysis | Unadjusted model |  | Adjusted model¶ |  |  | Descriptive analysis | Unadjusted model |  | Adjusted model¶ |  |
| Potential Risk factors | n/N(%) | OR (95%CI) | P-value* | AOR (95%CI) | P-value* | Potential Risk factors | n/N(%) | OR (95%CI) | P-value* | AOR (95%CI) | P-value* |
| *Community* |  |  |  |  |  | *Community* |  |  |  |  |  |
| *Z8* | 1609/46995(3.4) | Reference | <0.001 | Reference | <0.001 | *SA16* | 1379/27923(4.9) | Reference | <0.001 | Reference | <0.001 |
| *Z1* | 593/23096(2.6) | 0.73(0.58,0.91) |  | 0.77(0.62,0.97) |  | *SA13* | 591/15829(3.7) | 0.78(0.64,0.94) |  | 0.75(0.62,0.92) |  |
| *Z2* | 437/10615(4.1) | 1.22(0.91,1.62) |  | 1.33(1.00,1.78) |  | *SA14* | 344/7218(4.8) | 1.04(0.80,1.34) |  | 1.00(0.77,1.29) |  |
| *Z5* | 449/22942(2.0) | 0.56(0.44,0.70) |  | 0.54(0.43,0.68) |  | *SA18* | 1342/15707(8.5) | 1.90(1.57,2.29) |  | 1.80(1.49,2.16) |  |
| *Z6* | 327/16185(2.0) | 0.58(0.45,0.74) |  | 0.51(0.40,0.66) |  | *SA19* | 452/6779(6.7) | 1.42(1.11,1.80) |  | 1.21(0.95,1.54) |  |
| *Z9* | 1168/44068(2.7) | 0.74(0.63,0.88) |  | 0.78(0.66,0.92) |  | *SA20* | 755/10231(7.4) | 1.62(1.30,2.01) |  | 1.48(1.19,1.83) |  |
| *Z10* | 368/15828(2.3) | 0.66(0.52,0.84) |  | 0.66(0.52,0.84) |  |  | - | - | - | - | - |
| *Z11* | 332/14215(2.3) | 0.66(0.52,0.85) |  | 0.69(0.54,0.89) |  |  | - | - | - | - | - |
| *Age (in years)* |  |  |  |  |  | *Age (in years)* |  |  |  |  |  |
| *15-17* | 245/18615(1.3) | 0.70(0.6,0.80) | <0.001 | 0.77(0.67,0.89) | <0.001 | *15-17* | 196/5687(3.4) | 0.73(0.62,0.86) | <0.001 | 0.72(0.61,0.85) | <0.001 |
| *18-19* | 268/16443(1.6) | 0.85(0.74,0.98) |  | 0.90(0.78,1.03) |  | *18-19* | 199/4653(4.3) | 0.93(0.79,1.10) |  | 0.92(0.78,1.09) |  |
| *20-24* | 812/42393(1.9) | Reference |  | Reference |  | *20-24* | 644/14317(4.5) | Reference |  | Reference |  |
| *25-29* | 740/31618(2.3) | 1.22(1.10,1.35) |  | 1.14(1.03,1.26) |  | *25-29* | 730/13800(5.3) | 1.20(1.07,1.34) |  | 1.24(1.11,1.39) |  |
| *30-39* | 1362/42243(3.2) | 1.72(1.58,1.88) |  | 1.47(1.34,1.61) |  | *30-39* | 1223/21245(5.8) | 1.33(1.21,1.47) |  | 1.39(1.26,1.54) |  |
| *40-49* | 918/21663(4.2) | 2.31(2.10,2.54) |  | 1.91(1.72,2.11) |  | *40-49* | 903/12477(7.2) | 1.66(1.49,1.84) |  | 1.79(1.60,1.99) |  |
| *50+* | 938/20969(4.5) | 2.46(2.23,2.70) |  | 2.61(2.36,2.88) |  | *50+* | 968/11508(8.4) | 1.81(1.63,2.02) |  | 2.11(1.89,2.35) |  |
| *Sex* |  |  |  |  |  | *Sex* |  |  |  |  |  |
| *Male* | 2368/77959(3.0) | Reference | <0.001 | Reference | <0.001 | *Male* | 2628/34379(7.6) | Reference | <0.001 | Reference | <0.001 |
| *Female* | 2915/115985(2.5) | 0.82(0.78,0.87) |  | 0.74(0.70,0.79) |  | *Female* | 2235/49308(4.5) | 0.57(0.54,0.60) |  | 0.53(0.50,0.57) |  |
| **HIV status** |  |  |  |  |  | **HIV status** |  |  |  |  |  |
| *Tested HIV neg by CHiP* | 3087/133448(2.3) | Reference | <0.001 | Reference | <0.001 | *Tested HIV neg by CHiP* | 3261/47245(6.9) | Reference | <0.001 | Reference | <0.001 |
| *Self-reported HIV pos & NOT on ART* | 94/1758(5.3) | 2.49(2.01,3.08) |  | 2.26(1.83,2.81) |  | *Self-reported HIV pos & NOT on ART* | 106/753(14.1) | 2.34(1.90,2.90) |  | 2.48(2.00,3.09) |  |
| *self-reported HIV pos & on ART* | 1209/20078(6.0) | 2.76(2.58,2.96) |  | 2.28(2.12,2.46) |  | *self-reported HIV pos & on ART* | 550/8708(6.3) | 1.02(0.92,1.12) |  | 1.04(0.94,1.15) |  |
| *Tested HIV pos by CHiP* | 571/4380(13.0) | 6.51(5.91,7.17) |  | 5.92(5.36,6.55) |  | *Tested HIV pos by CHiP* | 188/1239(15.2) | 2.58(2.19,3.04) |  | 2.64(2.23,3.11) |  |
| *Not-tested/indeterminate* | 322/34280(0.9) | 0.39(0.35,0.44) |  | 0.35(0.31,0.40) |  | *Not-tested/indeterminate* | 758/25742(2.9) | 0.42(0.39,0.45) |  | 0.39(0.36,0.43) |  |
| **Household TB contact** |  |  |  |  |  | **Household TB contact** |  |  |  |  |  |
| *No* | 5085/192308(2.6) | Reference | <0.001 | Reference | <0.001 | *No* | 4611/83063(5.6) | Reference | <0.001 | Reference | <0.001 |
| *Yes* | 198/1636(12.1) | 4.99(4.28,5.82) |  | 4.52(3.85,5.30) |  | *Yes* | 252/624(40.4) | 10.13(8.55,11.98) |  | 10.27(8.63,12.23) |  |
| **Mobility** |  |  |  |  |  | **Mobility** |  |  |  |  |  |
| *Previously Resident* | 3485/135112(2.6) | Reference | <0.001 | Reference | <0.001 | *Previously Resident* | 3538/61813(5.7) | Reference | 0.013 | Reference | <0.001 |
| *Moved from within community* | 939/34069(2.8) | 1.09(1.01,1.17) |  | 1.18(1.09,1.28) |  | *Moved from within community* | 843/13378(6.3) | 1.12(1.03,1.22) |  | 1.16(1.07,1.26) |  |
| *Moved from outside community* | 859/24763(3.5) | 1.36(1.26,1.47) |  | 1.48(1.36,1.60) |  | *Moved from outside community* | 482/8496(5.7) | 1.11(1.00,1.23) |  | 1.14(1.02,1.26) |  |

Note: \* P-values from Likelihood ratio test; n/N(Yield) = newly diagnosed TB disease by CHIP/Number screened (TB disease yield expressed per 100,000
people screened); “-” Information missing; OR = Odds Ratio; AOR = Adjusted Odds Ratio; CI = Confidence Interval; CHiP = Community HIV care
Provider; pos = positive; neg = negative; ¶ Model adjusted for community, sex, age, HIV status, household TB contact and mobility

In R3, in both countries; female participants were less likely to have TB symptoms compared to males (Fig 2b, Table 1). Participants in older age-groups were more likely to have TB symptoms compared to younger age-groups. Participants who had moved were more likely to have TB symptoms compared to previous residents of the community (Table 1).

In Zambia in R3, participants newly diagnosed HIV-positive were more likely to have TB symptoms compared to those who tested HIV-negative, and those who self-reported being HIV-positive. However, the prevalence of TB symptoms was similar in those who self-reported being HIV-positive on ART and those who self-reported being HIV-positive but not on ART (Fig 2c, Table 1). Similar patterns were observed across R1-3 in Zambia, though in R1 and R2 the prevalence of TB symptoms was lower among individuals who self-reported being HIV-positive and on ART compared with those who self-reported being HIV-positive and not on ART (Fig 2c, S5 Fig).

In SA, participants newly diagnosed HIV-positive and participants who self-reported being HIV-positive and not on ART, were more likely to have TB symptoms compared to those who tested HIV-negative. However, there was no evidence of a difference in the prevalence of TB symptoms between those who tested HIV-negative and those who self-reported being HIV-positive and on ART. (Fig 2c, Table 1).

### Yield of newly diagnosed TB disease and factors associated with being newly diagnosed with TB disease

The yield of newly diagnosed TB disease per 100,000 people screened increased across R1-3 in Zambia [R1 = 80.7/100000, R2 = 92.8/100000 and R3 = 110.3/100000, p-value = 0.003] and was higher in SA compared to Zambia in R3 [Zambia:110/100,000 95% confidence interval (CI) (96, 126); SA: 380/100,000 95% CI (339, 424)] (Fig 2d).

Age, sex, household TB contact and HIV status in Zambia (R3) and community of residence, sex, HIV status and household TB contact in SA were associated with being newly diagnosed with TB disease (Table 2). In both countries, female participants were less likely to be newly diagnosed with TB disease compared to males [Zambia; Adjusted Odds Ratio (AOR) = 0.32 95% CI (0.24, 0.42) and SA; AOR = 0.45 95% CI (0.36, 0.56)] (Fig 2e, Table 2), while household TB contacts were more likely to be newly diagnosed with TB disease [Zambia; AOR = 7.44 95% CI (4.27, 12.95)] and SA; AOR = 13.40 95% CI(9.11, 19.70)] (Table 2).

**Table 2:**
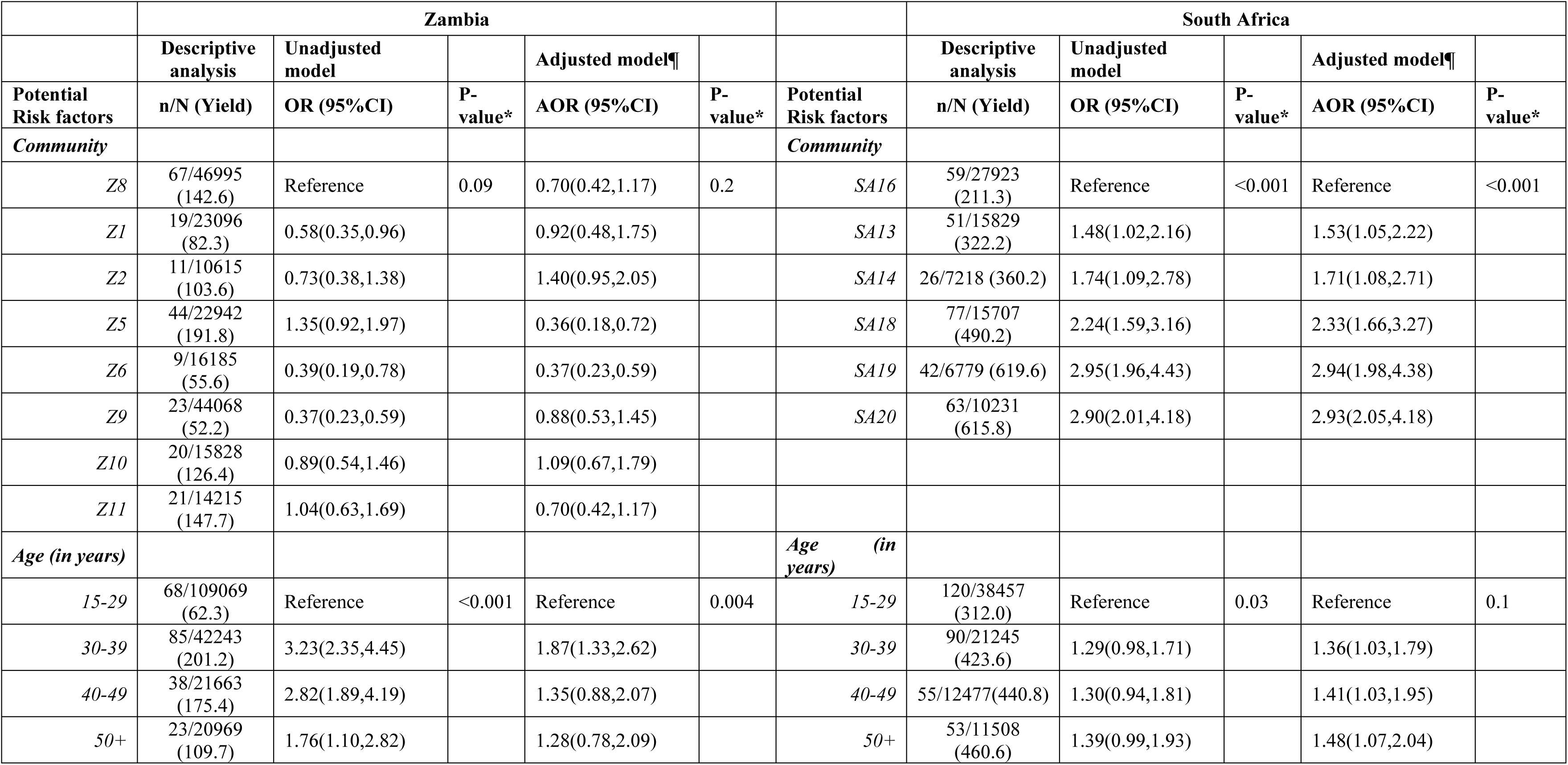
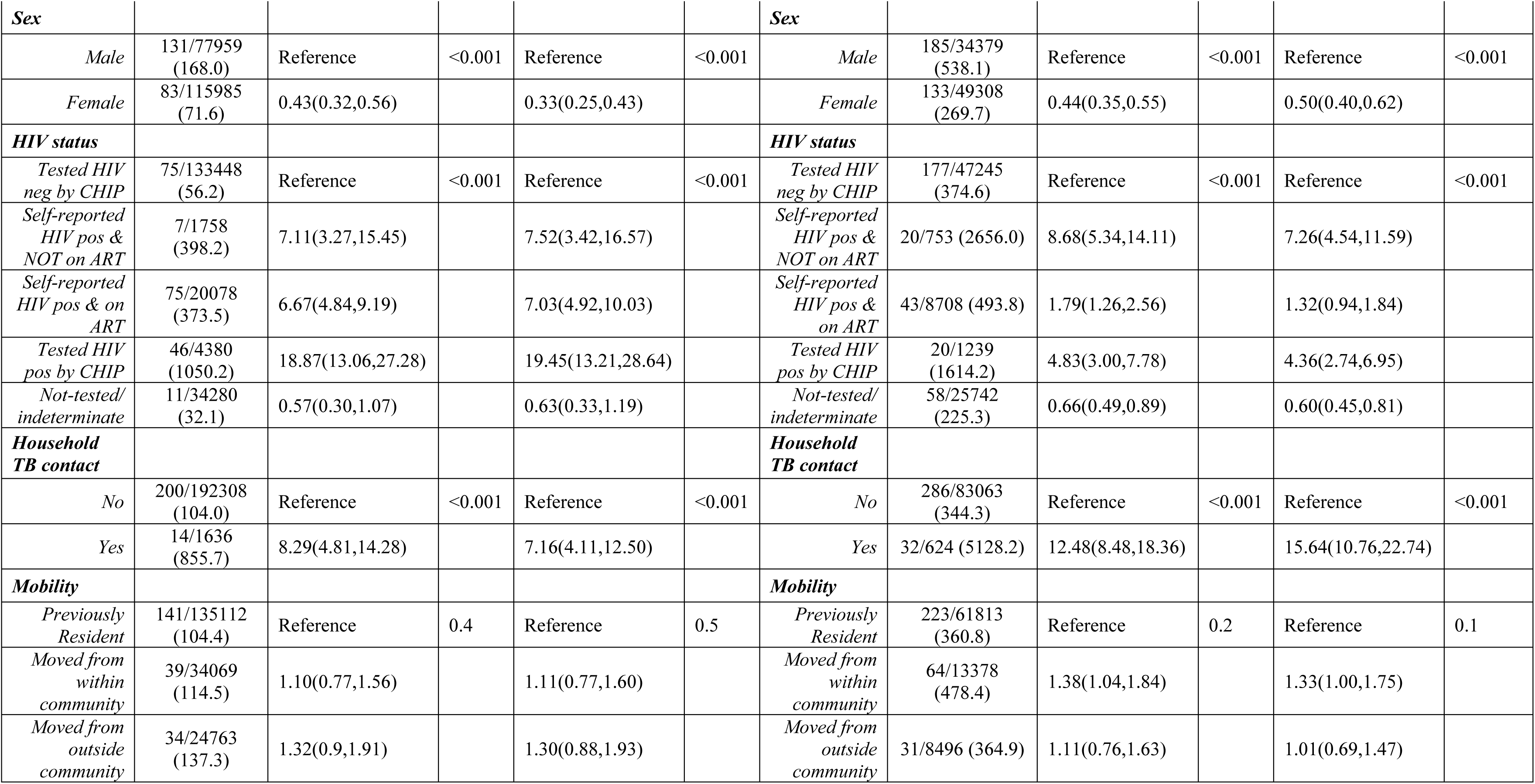
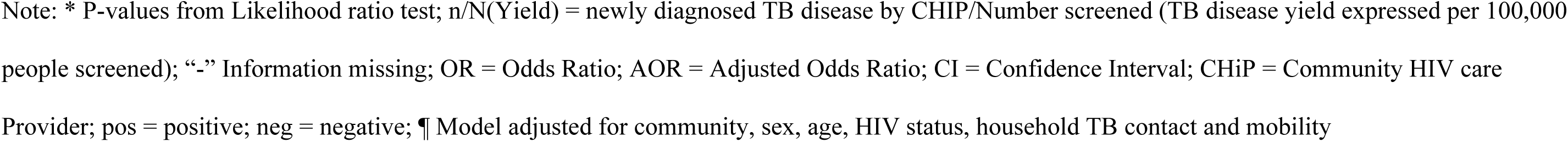
Factors associated with being newly diagnosed with TB Disease in Round 3 (October 2016 to December 2017) of the intervention in Zambia & South Africa.

|  | Zambia |  |  |  |  |  | South Africa |  |  |  |  |
| --- | --- | --- | --- | --- | --- | --- | --- | --- | --- | --- | --- |
|  | Descriptive analysis | Unadjusted model |  | Adjusted model¶ |  |  | Descriptive analysis | Unadjusted model |  | Adjusted model¶ |  |
| Potential Risk factors | n/N (Yield) | OR (95%CI) | P-value* | AOR (95%CI) | P-value* | Potential Risk factors | n/N (Yield) | OR (95%CI) | P-value* | AOR (95%CI) | P-value* |
| *Community* |  |  |  |  |  | *Community* |  |  |  |  |  |
| Z8 | 67/46995 (142.6) | Reference | 0.09 | 0.70(0.42,1.17) | 0.2 | SA16 | 59/27923 (211.3) | Reference | <0.001 | Reference | <0.001 |
| Z1 | 19/23096 (82.3) | 0.58(0.35,0.96) |  | 0.92(0.48,1.75) |  | SA13 | 51/15829 (322.2) | 1.48(1.02,2.16) |  | 1.53(1.05,2.22) |  |
| Z2 | 11/10615 (103.6) | 0.73(0.38,1.38) |  | 1.40(0.95,2.05) |  | SA14 | 26/7218 (360.2) | 1.74(1.09,2.78) |  | 1.71(1.08,2.71) |  |
| Z5 | 44/22942 (191.8) | 1.35(0.92,1.97) |  | 0.36(0.18,0.72) |  | SA18 | 77/15707 (490.2) | 2.24(1.59,3.16) |  | 2.33(1.66,3.27) |  |
| Z6 | 9/16185 (55.6) | 0.39(0.19,0.78) |  | 0.37(0.23,0.59) |  | SA19 | 42/6779 (619.6) | 2.95(1.96,4.43) |  | 2.94(1.98,4.38) |  |
| Z9 | 23/44068 (52.2) | 0.37(0.23,0.59) |  | 0.88(0.53,1.45) |  | SA20 | 63/10231 (615.8) | 2.90(2.01,4.18) |  | 2.93(2.05,4.18) |  |
| Z10 | 20/15828 (126.4) | 0.89(0.54,1.46) |  | 1.09(0.67,1.79) |  |  |  |  |  |  |  |
| Z11 | 21/14215 (147.7) | 1.04(0.63,1.69) |  | 0.70(0.42,1.17) |  |  |  |  |  |  |  |
| *Age (in years)* |  |  |  |  |  | *Age (in years)* |  |  |  |  |  |
| 15-29 | 68/109069 (62.3) | Reference | <0.001 | Reference | 0.004 | 15-29 | 120/38457 (312.0) | Reference | 0.03 | Reference | 0.1 |
| 30-39 | 85/42243 (201.2) | 3.23(2.35,4.45) |  | 1.87(1.33,2.62) |  | 30-39 | 90/21245 (423.6) | 1.29(0.98,1.71) |  | 1.36(1.03,1.79) |  |
| 40-49 | 38/21663 (175.4) | 2.82(1.89,4.19) |  | 1.35(0.88,2.07) |  | 40-49 | 55/12477(440.8) | 1.30(0.94,1.81) |  | 1.41(1.03,1.95) |  |
| 50+ | 23/20969 (109.7) | 1.76(1.10,2.82) |  | 1.28(0.78,2.09) |  | 50+ | 53/11508 (460.6) | 1.39(0.99,1.93) |  | 1.48(1.07,2.04) |  |
| **Sex** |  |  |  |  |  | **Sex** |  |  |  |  |  |
| *Male* | 131/77959<br>(168.0) | Reference | <0.001 | Reference | <0.001 | *Male* | 185/34379<br>(538.1) | Reference | <0.001 | Reference | <0.001 |
| *Female* | 83/115985<br>(71.6) | 0.43(0.32,0.56) |  | 0.33(0.25,0.43) |  | *Female* | 133/49308<br>(269.7) | 0.44(0.35,0.55) |  | 0.50(0.40,0.62) |  |
| **HIV status** |  |  |  |  |  | **HIV status** |  |  |  |  |  |
| *Tested HIV*<br>*neg by CHIP* | 75/133448<br>(56.2) | Reference | <0.001 | Reference | <0.001 | *Tested HIV*<br>*neg by CHIP* | 177/47245<br>(374.6) | Reference | <0.001 | Reference | <0.001 |
| *Self-reported*<br>*HIV pos &*<br>*NOT on ART* | 7/1758<br>(398.2) | 7.11(3.27,15.45) |  | 7.52(3.42,16.57) |  | *Self-reported*<br>*HIV pos &*<br>*NOT on ART* | 20/753 (2656.0) | 8.68(5.34,14.11) |  | 7.26(4.54,11.59) |  |
| *Self-reported*<br>*HIV pos & on*<br>*ART* | 75/20078<br>(373.5) | 6.67(4.84,9.19) |  | 7.03(4.92,10.03) |  | *Self-reported*<br>*HIV pos &*<br>*on ART* | 43/8708 (493.8) | 1.79(1.26,2.56) |  | 1.32(0.94,1.84) |  |
| *Tested HIV*<br>*pos by CHIP* | 46/4380<br>(1050.2) | 18.87(13.06,27.28) |  | 19.45(13.21,28.64) |  | *Tested HIV*<br>*pos by CHIP* | 20/1239<br>(1614.2) | 4.83(3.00,7.78) |  | 4.36(2.74,6.95) |  |
| *Not-tested/*<br>*indeterminate* | 11/34280<br>(32.1) | 0.57(0.30,1.07) |  | 0.63(0.33,1.19) |  | *Not-tested/*<br>*indeterminate* | 58/25742<br>(225.3) | 0.66(0.49,0.89) |  | 0.60(0.45,0.81) |  |
| **Household**<br>**TB contact** |  |  |  |  |  | **Household**<br>**TB contact** |  |  |  |  |  |
| *No* | 200/192308<br>(104.0) | Reference | <0.001 | Reference | <0.001 | *No* | 286/83063<br>(344.3) | Reference | <0.001 | Reference | <0.001 |
| *Yes* | 14/1636<br>(855.7) | 8.29(4.81,14.28) |  | 7.16(4.11,12.50) |  | *Yes* | 32/624 (5128.2) | 12.48(8.48,18.36) |  | 15.64(10.76,22.74) |  |
| **Mobility** |  |  |  |  |  | **Mobility** |  |  |  |  |  |
| *Previously*<br>*Resident* | 141/135112<br>(104.4) | Reference | 0.4 | Reference | 0.5 | *Previously*<br>*Resident* | 223/61813<br>(360.8) | Reference | 0.2 | Reference | 0.1 |
| *Moved from*<br>*within*<br>*community* | 39/34069<br>(114.5) | 1.10(0.77,1.56) |  | 1.11(0.77,1.60) |  | *Moved from*<br>*within*<br>*community* | 64/13378<br>(478.4) | 1.38(1.04,1.84) |  | 1.33(1.00,1.75) |  |
| *Moved from*<br>*outside*<br>*community* | 34/24763<br>(137.3) | 1.32(0.9,1.91) |  | 1.30(0.88,1.93) |  | *Moved from*<br>*outside*<br>*community* | 31/8496 (364.9) | 1.11(0.76,1.63) |  | 1.01(0.69,1.47) |  |

Note: \* P-values from Likelihood ratio test; n/N(Yield) = newly diagnosed TB disease by CHIP/Number screened (TB disease yield expressed per 100,000 people screened); “-” Information missing; OR = Odds Ratio; AOR = Adjusted Odds Ratio; CI = Confidence Interval; CHiP = Community HIV care Provider; pos = positive; neg = negative; ¶ Model adjusted for community, sex, age, HIV status, household TB contact and mobility

In R3 Zambia, PLHIV were more likely to be newly diagnosed with TB disease [i.e. self-reported HIV-positive not on ART: AOR = 7.52 95% CI (3.42, 16.57); Self-reported HIV-positive on ART: AOR = 7.03 95%CI (4.92, 10.03) and newly diagnosed HIV-positive: AOR = 19.45 95%CI (13.21, 28.64)] compared to participants who tested HIV-negative. In R3 SA, participants who self-reported being HIV-positive and not on ART [AOR = 7.26 95% CI (4.54, 11.59)] and participants newly diagnosed HIV-positive were more likely to be newly diagnosed with TB disease [AOR = 4.36 95%CI (2.74, 6.95)] compared to participants who tested HIV-negative. However, there was no clear difference between participants who tested HIV-negative and those who self-reported being HIV-positive on ART (Fig 2f, Table 2). Similar patterns were observed across R1-3 in Zambia (S6 Fig) Across R1-3 in Zambia an average of 80% of participants diagnosed with TB self-reported to have started treatment, and 92% in R3 in SA, (Fig 1), a higher proportion being seen in males in R1-2 Zambia and R3 SA (S3 Table).

### Association between repeat screening and the yield of TB disease

Among the 193,944 participants screened in R3 Zambia, 84,677 (43.7%) were being screened for the first time, 55,616 (28.9%) had been screened once before and 53,651 (27.7%) had been screened twice before (Fig 3). The yield decreased (p-value (test for trend) = 0.011) with increasing repeat TB screenings. Participants screened twice before were less likely to be newly diagnosed with TB disease compared to those screened for the first time [AOR = 0.62 95% CI (0.43, 0.90)] (Table 3). The findings are similar when restricted to participants resident across all the three rounds in Zambia (S7 Fig, S4 Table).

**Figure 3:**
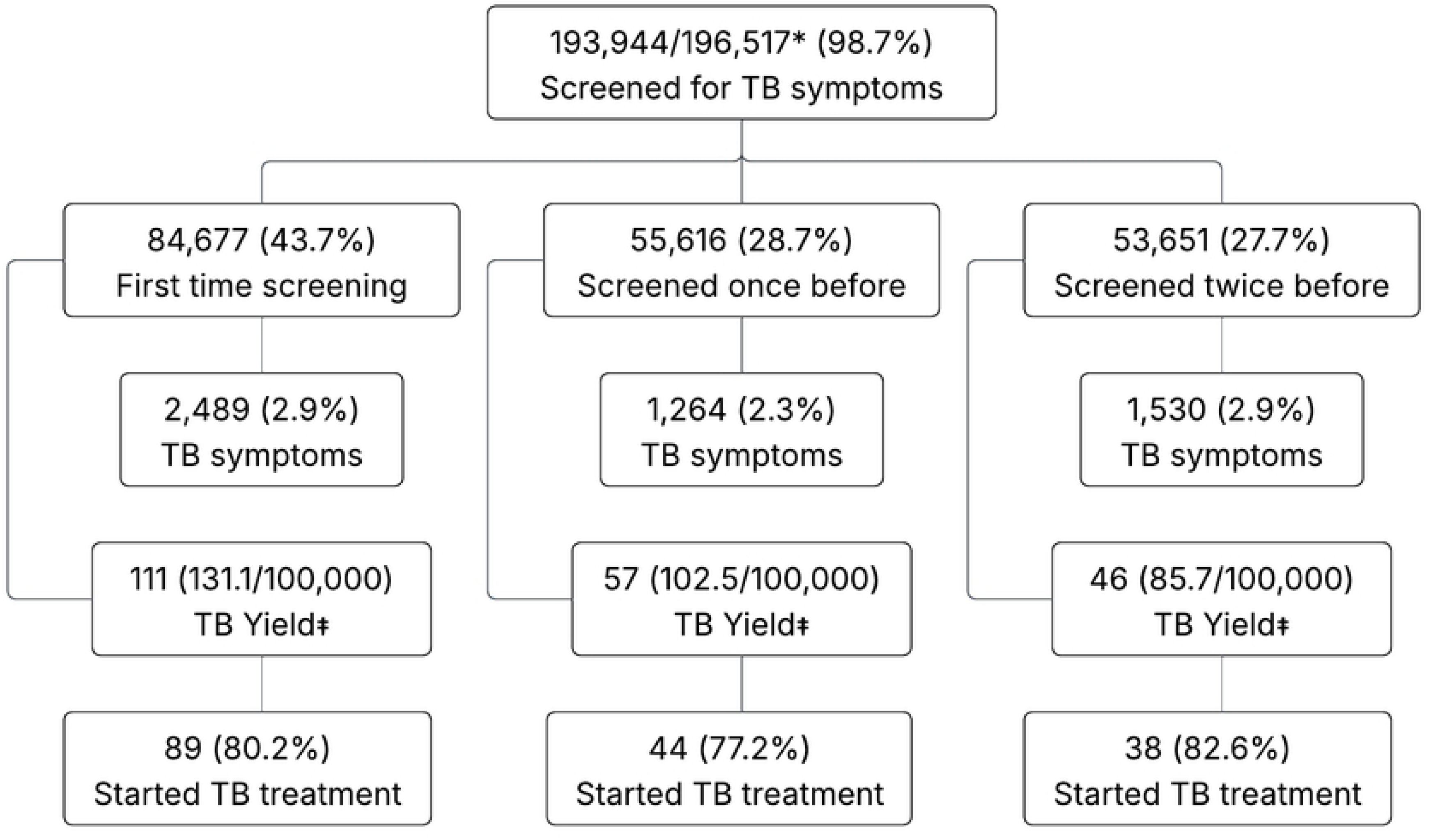
TB outcomes stratified by different screening patterns, among participants screened for TB symptoms in Round 3 of the Intervention in Zambia. Note: * Among all those who participated in R3 in Zambia; ‡ TB yield (per 100,000 people screened) denominator is the number screened for TB symptoms

**Table 3:** Yield of newly diagnosed TB disease stratified by screening patterns, during the third round of population-wide screening in 2017, in a population in which 2 previous rounds of population-wide screening were conducted during 2014-2016 in Zambia.

|  | Descriptive analysis | Unadjusted model |  | Adjusted model¶ |  |
| --- | --- | --- | --- | --- | --- |
|  | n/N(Yield) | OR (95%CI) | p-value* | AOR (95%CI) | p-value* |
| **Screening‡** |  |  |  |  |  |
| *First time screening* | 111/84677 (131.1) | Reference | 0.036 | Reference | 0.039 |
| *Screened once before* | 57/55616 (102.5) | 0.78 (0.57,1.08) |  | 0.86 (0.62,1.20) |  |
| *Screened twice before* | 46/53651 (85.7) | 0.65 (0.46,0.92) |  | 0.62 (0.43,0.90) |  |

**Note:** \* P-values from Likelihood ratio test; ‡ Number of times screened in the CHiP zone where an individual was resident in R3; n/N(Yield) = newly diagnosed TB disease by CHIP/Number screened (TB disease yield expressed per 100,000 people screened); OR = Odds Ratio; AOR = Adjusted Odds Ratio; CI = Confidence Interval; CHiP = Community HIV care Provider; pos = positive; neg = negative; ¶ Model adjusted for triplet, sex, age, HIV-status and household TB contact

## Discussion

Our results demonstrated TB symptom screening can be integrated into a universal HIV testing programme. For a trial of such magnitude, PopART achieved high community coverage, with participation rates of 83%, 71% and 75% across R1-3 in Zambia and 63% in SA in R3.

The yield of newly diagnosed TB disease was higher in R3 SA (380/100000) than in R3 Zambia (110/100000) study communities, reflecting expected TB burden differences by country. Yield also increased across intervention rounds in Zambia. While this finding might seem unexpected, two reasons could explain this: One, the primary objective of PopART was to deliver UTT for HIV, so CHiPs may have initially focused more on the HIV components of the intervention, particularly in R1, with a growing emphasis on TB screening in later rounds, as indicated by the increase in the prevalence of self-reported TB symptoms. Two, participants may have become more open to discussing TB as they better understood its importance. Study teams noted the unexpectedly low symptom prevalence in R1 compared to other studies in similar settings, and during R2-R3 made concerted efforts to train staff and educate communities on TB’s significance.

A recent TB prevalence survey (2019–2021) estimated TB prevalence (per 100,000) at 500 Zambia and 1,600 in SA [15]. Screening included both symptom enquiry (i.e., cough or weight loss ≥1 month; night sweats, chest pain, or fever ≥2 weeks) and chest X-ray. Symptoms were present in 3.8% of participants in Zambia and 4.4% in SA, but only 42% and 20% of bacteriologically confirmed TB cases screened positive for symptoms alone. This suggests that the symptom-based approach used by CHiPs likely identified only 20–25% of individuals with prevalent TB, a disappointingly low proportion. This highlights a key limitation of symptom-only screening and the need for more sensitive tools such as chest X-ray [2,16]. Notably, our screening included only cough, weight loss, and night sweats, possibly further reducing sensitivity. In Zambia, diagnostics followed national guidelines using both Xpert and smear microscopy, the latter of which has lower sensitivity [17,18]. As a result, the PopART intervention likely detected only a minority of prevalent TB cases and had no measurable impact on TB prevalence compared to the control arm [15].

In contrast, the ACT3 trial in Vietnam, using repeated universal Xpert testing irrespective of symptoms, demonstrated a 44% reduction in TB prevalence [3]. This approach involved repeated rounds of Xpert MTB/RIF testing for all individuals, regardless of symptoms, over three years. While such an intensive strategy is likely unfeasible in most countries, it underscores the need to expand beyond symptom-based screening in high-burden settings.

In R3 Zambia, we showed that among a population repeatedly visited and screened for TB symptoms, the odds of being newly diagnosed with TB were 38% lower after the first year among those screened twice before, compared to those screened for the first time. Similarly, in the ACT3 trial, a reduction in prevalent TB observed in the fourth year following three years of active community-wide screening, suggests that repeated screening may help reduce TB prevalence [3]. A recent systematic review also suggests annual community-wide screening for pulmonary TB can reduce prevalence, provided it uses accurate tests and achieves high coverage [19]. In our study, a surprisingly high proportion of participants (44%) were screened for the first time in the zone where they resided during R3, despite relatively high screening coverage per round. This highlights the high population mobility, which may have diluted the estimated effects of repeated screening.

Consistent with previous studies, we found the highest yield of undiagnosed TB among individuals newly diagnosed with HIV, followed by self-reported being HIV-positive, and the lowest yield among HIV-negative individuals [20–22]. By R3, a high proportion of self-reported HIV-positive individuals were on ART [14], which may explain their lower TB yield compared to those newly diagnosed. This pattern was more pronounced in Zambia than SA, possibly reflecting differences in ART coverage or timing of HIV acquisition among those not yet on treatment. Also consistent with previous studies, men - especially older men - and household TB contacts had higher rates of undiagnosed TB [8,23,24]. Yield was lowest in those aged 15-29 years, peaked at 30-39 years, and declined among older adults in Zambia, while remaining high among older adults in SA. These age and sex patterns align with recent prevalence surveys in both countries [20,22].

A key strength of this study is its integration within the large PopART trial, which achieved high household coverage, substantial sample size, and strong individual participation across annual rounds in communities with high TB/HIV burden. The ability to revisit the same communities annually with a door-to-door TB/HIV intervention - delivered by trained community health workers from within those communities - enabled a robust, real-world assessment of TB symptom screening over time. These findings are likely generalisable to other urban Southern Africa settings with similarly high TB/HIV prevalence and mobility.

A key limitation is probable under-ascertainment of TB due to reliance on symptom-based screening, which misses individuals without typical TB symptoms [23,25]. In Zambia, the use of smear microscopy, less sensitive than Xpert or culture, may have further limited case detection [17]. Lower male participation (63% vs 85% in women in R3 Zambia; 55% vs 70% overall) may have led to underestimation of TB burden in this higher-risk group.

Nonetheless, our study shows that, while community-wide TB symptom screening - combined with smear microscopy or a rapid nucleic acid amplification test (Xpert MTB/RIF, Cepheid) – can identify undiagnosed TB, integration with HIV screening may result in lesser emphasis on TB screening and suboptimal identification of prevalent TB. Including chest X-rays in TB screening, known to increase diagnostic yield, would be valuable [15,16,26]. The advent of ultra-portable X-ray systems fitted with computer-aided detection (CAD) software since 2018-2019 has made TB symptom enquiry increasingly scalable [27,28]. These technologies offer enhanced potential for community-based systematic screening by improving sensitivity and operational efficiency in high-burden settings [29].

In conclusion, our findings show that repeated community-wide TB symptom screening, delivered alongside universal HIV testing, identified undiagnosed TB disease, particularly among men, PLHIV, and those not previously screened. However, the yield was relatively low compared to estimated TB prevalence, highlighting the limitations of symptom-based screening and the likelihood that many individuals with prevalent TB were missed.

## Data Availability

We will provide aggregate data sufficient to replicate the main analyses presented in this paper. These aggregate data will include counts of tuberculosis screening and diagnostic outcomes stratified by country, round, trial arm, community, gender, age group, HIV status, and participation in previous screening rounds. The protocol approved by the ethics committees specifies that intervention data can only be shared in aggregate form. For queries related to these data, the PI of the HPTN Statistical Data Management Centre (SDMC), Deborah Donnell, can be contacted.

## Acknowledgments

We thank all members of the HPTN 071 (PopART) Study Team and all study participants and their communities, for their contributions to this research.

## Supporting information

**S1 STROBE Checklist. STROBE, strengthening the reporting of observational studies in epidemiology.**

**S1 Table: Household census, enumeration and participation eligibility by age and sex across R1-3 in Zambia and R3 in South Africa.** Note: * ≥18-year-olds in R1 and ≥15-year-olds in R2-3; %(n/N) = Percent (number/denominator); R1-3 = Round 1-3 (January 2014 – December 2017); R3 = Round 3 (October 2016 – December 2017); “-” Information missing; ART = Antiretroviral Therapy; HIV = Human Immunodeficiency Virus; CHiP = Community HIV care Provider; pos = positive; neg = negative.

**S2 Table: Participation patterns by age, sex and HIV status across R1-3 in Zambia and R3 in South Africa.** Note: * ≥18-year-olds in R1 and ≥15-year-olds in R2-3; R1-3 = Round 1-3 (January 2014 – December 2017); R3 = Round 3 (October 2016 – December 2017); %(n/N) = Percent (number/total number); “-” Information missing; ART = Antiretroviral Therapy; HIV = Human Immunodeficiency Virus; CHiP = Community HIV care Provider; pos = positive; neg = negative.

**S1 Fig: Participation trends by age and sex across R1-3 in Zambia and R3 in South Africa**. Note: R1-3 = Round 1-3 (January 2014 – December 2017); R3 = Round 3 (October 2016 – December 2017).

**S2 Fig: Patterns of the prevalence of TB symptoms across intervention rounds and communities in Zambia.**

**S3 Fig: Patterns of the prevalence of TB symptoms across intervention rounds and communities in Zambia (separately for males and females).**

**S4 Fig: Patterns of the prevalence of TB symptoms across intervention rounds and communities in Zambia (separately by age groups i.e.15-29, 30-39, 20-49 and 50+ years).**

**S5 Fig: Factors associated with having TB symptoms across Round 1-3 in Zambia only.** Note: AOR = Adjusted Odds Ratio (adjusted for community, sex, age, HIV-status, household TB contact and mobility). R1 – 3 = Round 1-3 between January 2014 – December 2017.

**S6 Fig: Factors associated with being newly diagnosed with TB Disease across Round 1-3 in Zambia only.** Note: R1-3 = Round 1-3 between January 2014 – December 2017.

**S3 Table: TB symptom screening, sputum collection and recording of results, patterns by age, sex and HIV status across R1-3 in Zambia and R3 in South Africa.** Note: * ≥18-year-olds in R1 and ≥15-year-olds in R2-3; %(n/N) = Percent (number/total number); R1-3 = Round 1-3 (January 2014 – December 2017); R3 = Round 3 (October 2016 – December 2017); “-” Information missing; ART = Antiretroviral Therapy; HIV = Human Immunodeficiency Virus; CHiP = Community HIV care Provider; pos = positive; neg = negative; ‡ For every 100,000 people screened.

**S4 Table: Yield of newly diagnosed TB disease stratified by screening patterns, during the third round of population-wide screening in** 2017**, only for the population resident across R1 to R3 (**2014**-17) in Zambia.** Note:* P-values from Likelihood ratio test; ‡ Number of times screened in the CHiP zone where an individual was resident in R3; n/N(Yield) = newly diagnosed TB disease by CHIP/Number screened (TB disease yield expressed per 100,000 people screened); OR = Odds Ratio; AOR = Adjusted Odds Ratio; CI = Confidence Interval; CHiP = Community HIV care Provider; pos = positive; neg = negative; ¶ Model adjusted for triplet, sex, age, HIV-status and household TB contact.

**S7 Fig: TB outcomes stratified by different screening patterns, among participants screened for TB symptoms in Round 3 of the Intervention in Zambia and were residents across R1 to R3 (**2014**-17) in Zambia.** Note: * Among all those who participated in R3 in Zambia; ‡ TB yield (per 100,000 people screened) denominator is the number screened for TB symptoms.

